# Polymerized type I collagen down-regulates STAT-1 phosphorylation through engagement to LAIR-1 in M1-macrophages avoiding long COVID

**DOI:** 10.1101/2023.07.01.23292108

**Authors:** Elizabeth Olivares-Martínez, Diego Francisco Hernández-Ramírez, Carlos Alberto Núñez-Álvarez, Mónica Chapa-Ibarguengoitia, Silvia Méndez-Flores, Ángel Priego- Ranero, Daniel Azamar-Llamas, Héctor Olvera-Prado, Kenia Ilian Rivas-Redonda, Eric Ochoa-Hein, Luis Gerardo López-Mosqueda, Estefano Rojas-Castañeda, Said Urbina-Terán, Luis Septién-Stute, Thierry Hernández-Gilsoul, Diana Aguilar-León, Gonzalo Torres-Villalobos, Janette Furuzawa-Carballeda

**Affiliations:** Department of Immunology and Rheumatology, Instituto Nacional de Ciencias Médicas y Nutrición Salvador Zubirán, Mexico City, Mexico; Department of Radiology, Instituto Nacional de Ciencias Médicas y Nutrición Salvador Zubirán, Mexico City, Mexico; Department of Dermatology, Instituto Nacional de Ciencias Médicas y Nutrición Salvador Zubirán, Mexico City, Mexico; Department of Internal Medicine, Instituto Nacional de Ciencias Médicas y Nutrición Salvador Zubirán, Mexico City, Mexico; Department of Anesthesiology, Instituto Nacional de Ciencias Médicas y Nutrición Salvador Zubirán, Mexico City, Mexico; Department of Hospital Epidemiology, Instituto Nacional de Ciencias Médicas y Nutrición Salvador Zubirán, Mexico City, Mexico; Emergency Department, Instituto Nacional de Ciencias Médicas y Nutrición Salvador Zubirán, Mexico City, Mexico; Department of Pneumology, Instituto Nacional de Ciencias Médicas y Nutrición Salvador Zubirán, Mexico City, Mexico; Department of Pathology, Instituto Nacional de Ciencias Médicas y Nutrición Salvador Zubirán, Mexico City, Mexico; Departments of Experimental Surgery and Surgery, Instituto Nacional de Ciencias Médicas y Nutrición Salvador Zubirán, Mexico City, Mexico

## Abstract

**Background:** The polymerized type I collagen (PTIC) is a γ-irradiated mixture of pepsinized porcine type I collagen and polyvinylpyrrolidone (PVP). It has immunomodulatory properties. However, the receptor and signaling pathway through which it exerts its therapeutic effects has not yet been identified.

**Aim:** To evaluate LAIR-1 as a potential receptor for PTIC and the signaling pathway evoked by ligand-receptor binding.

**Methods:** LAIR-1 binding assay was performed by incubating various concentrations of recombinant human LAIR-1 with native type I collagen or PTIC. Macrophages M1- derived from THP-1 cells were cultured with 2-10% PTIC for 24 h. Cell lysates from THP- 1, monocytes-like cells (MLCs), M1, M1+IFN-γ, M1+LPS, and 2 or 10% PTIC treated M1 were analyzed by western blot for the transcription factors NF-κB (p65), p38, STAT-1, and pSTAT-1. Cytokines, Th1 cells, and M1/M2 macrophages were analyzed by luminometry and flow cytometry from blood samples of symptomatic COVID-19 outpatients on treatment with intramuscular administration of PTIC.

**Results:** PTIC binds LAIR-1 with a similar affinity to native collagen. This binding decreases STAT-1 signaling IFN-γ-induced and IL-1β expression in M1 macrophages by down-regulating STAT-1 phosphorylation. Moreover, intramuscular PTIC treatment of symptomatic COVID-19 outpatients decreased at statistically significant levels the percentage of M1 macrophages and cytokines (IP-10, MIF, eotaxin, IL-8, IL-1RA, and M-CSF) associated with STAT-1 transcription factor and increased M2 macrophages and Th1 cells. The downregulation of inflammatory mediators was related to better oxygen saturation and decreased dyspnea, chest pain, cough, and chronic fatigue syndrome in the acute phase of infection and the long term.

**Conclusion:** PTIC is an agonist of LAIR-1 and down-regulates STAT-1 phosphorylation. PTIC could be relevant for treating STAT-1-mediated inflammatory diseases, including COVID-19 and long COVID

## 1 INTRODUCTION

Polymerized type I collagen (PTIC) is a γ-irradiated mixture of pepsinized porcine type I collagen and polyvinylpyrrolidone (PVP) in a citrate buffer solution. At 37°C and neutral pH, the molecule does not form a gel, like collagen does, and its electrophoretic, physicochemical, and pharmacological properties are modified by the covalent bond between the protein and the PVP moiety.^1^

Previous works have demonstrated that PTIC has immunomodulatory properties. The addition of 1% PTIC to synovial tissue cultures from patients with rheumatoid arthritis or osteoarthritis downregulates the following: proinflammatory cytokines (IL-1β, TNF-α, IL-8, IL- 17, IFN-γ, PDGF, and TGF-β1); the expression of adhesion molecules such as endothelial leukocyte adhesion molecule-1 (ELAM-1), vascular cell adhesion molecule-1 (VCAM-1) and intercellular adhesion molecule-1 (ICAM-1); the expression of cyclooxygenase (Cox)-1 enzyme; and the collagenolytic activity. Moreover, PTIC has been shown to induce a positive regulation over the tissue inhibitor of metalloproteases-1 (TIMP-1), the production of IL-10, and the presence of regulatory T cells.^1–14^

Studies of intramuscular PTIC administration to patients with moderate-severe COVID-19 were associated with the down-regulation of the hyperinflammatory syndrome and better oxygen saturation values compared to placebo. Also, PTIC shortened symptom duration. One day after the last administration of PTIC, a higher mean oxygen saturation value and a higher proportion of patients retaining oxygen saturation values ≥92% were observed. This could be related to a decrease in dyspnea and chest pain, as well as cough. An unadjusted accelerated failure time model showed that the PTIC group achieved the outcome 2.70-fold faster (P<0.0001) than the placebo. Symptom duration in the PTIC group was reduced by 6.1±3.2 days vs. placebo. No differences in adverse effects were observed between the groups.^15–18^

To date, neither the receptor nor the signaling pathway of PTIC has been described. Thus, leukocyte-associated immunoglobulin-like receptor1 (LAIR-1 o CD305) was evaluated as a potential receptor for PTIC. LAIR-1 is a transmembrane inhibitory receptor that contains two immunoreceptors tyrosine-based inhibitory motif (ITIM) domains in its cytoplasmatic region.^19^ Collagen is a natural ligand for LAIR-1, its engagement on immune cells downregulates excessive inflammation.^20^ LAIR-1 is expressed in most hematopoietic cells, including T and B cells, neutrophils, dendritic cells, natural killers, monocyte, and macrophages.^21^ In this study, we show evidence of LAIR as one possible receptor for PTIC through *in vitro* analysis of THP-1 cells polarized to M1 macrophages treated with PTIC. PTIC binds LAIR-1 with a similar affinity to native collagen. This binding decreases the signal transducer and activator of transcription 1 (STAT-1), signaling IFN-γ-induced and IL- 1β expression in M1 macrophages by downregulating STAT-1 phosphorylation. In hyperinflammatory syndromes like COVID-19, PTIC administration induces M1 to M2 polarization and the decrease of chemokines and growth factors associated with STAT-1 transcription factor improving the acute phase of the infection and avoiding de long COVID.

## 2 MATERIALS AND METHODS

### 2.1 Cell Culture

Human monocytic THP-1 was maintained in culture with GIBCO RPMI 1640 (ThermoFisher Sci. USA) and 10% heat-inactivated fetal bovine serum (PAN-Biotech, De) at 37 °C, with 5% CO2 and 95% relative humidity.

### 2.2 Cell differentiation and treatments

The THP-1 cells were differentiated into macrophage-like cells (MLCs) by 72 h incubation with 100 nM phorbol-12-myristate13-acetate (PMA, Sigma P8139). Then MLCs were polarized in M1-macrophages by incubation with 20 ng/ml of IFN-γ and 1μg/ml of LPS for 24 h.^22^ M1-macrophages were cultured with different concentrations PTIC (2, 5, and 10 %), anti-LAIR-1 (1:100 dilution), (HycultBiotech # HM2364-100UG), or anti-LAIR-1 (1:100 dilution) + PTIC (10%) for 24 h at 37°C, with 5% CO2 and 95% relative humidity.

### 2.3 Flow cytometry

The MLCs and macrophages-M1 were incubated with or without treatment, with 5 μL of Human TruStain FcXTM (BioLegend Inc.) per million cells in 100 μl PBS for 10 minutes. Then they were labeled with 3 µL of anti-human: (a) CD36 FITC, CD16 PeCy, and CD86 APC or (b) CD14 FITC, CD16 PeCy, and CD163 APC antibodies in separated tubes for 20 min at room temperature in the dark. Cells were permeabilized with 200 µL of cytofix/cytoperm solution (BD Biosciences) at 4°C for 30 min. Intracellular staining was performed with an anti-human: (a) IL-1β PE or (b) IL-10 PE-labeled mouse monoclonal antibodies for 30 min at 4°C in the dark. An electronic gate was made for live cells (FCSA vs. FCSH), then for (a) CD16^+^/CD36^+^/CD86^+^ and (b) CD14^+^/CD16^hi^/CD163^+^ cells. Results are expressed as the relative percentage of (a) IL-1β^+^ and (b) IL-10^+^-expressing cells in each gate. As isotype control, IgG1-FITC/IgG1-PE/CD45-PeCy5 mouse IgG1 *kappa* (BD Tritest, BD Biosciences) was used to set the threshold and gates in the cytometer. We ran an unstained (autofluorescence control) and permeabilized PBMC sample. Autofluorescence control was compared to single-stained cell positive controls to confirm that the stained cells were on the scale for each parameter. Besides, BD Calibrate 3 beads were used to adjust instrument settings, set fluorescence compensation, and check instrument sensitivity (BD calibrates, BD Biosciences). Fluorescence minus one (FMO) controls were stained in parallel using the panel of antibodies with the sequential omission of intracellular antibodies. Finally, cell subsets were analyzed by flow cytometry with an Accuri C6 (BD Biosciences) in a blind manner regarding the clinical classification of the sample. Events (50,000–100,000) were recorded for each sample and analyzed with the FlowJo X software (Tree Star, Inc.).

### 2.4 Western blotting

Whole-cell lysates were generated in RIPA lysis buffer with 1mM phenylmethylsulfonyl fluoride (PMSF) and incubated for 15 min, 4°C. The supernatant was collected after centrifugation (13000 rpm, 15 min, four °C). Protein concentration was determined with a bicinchoninic acid assay. The protein solutions were loaded onto SDS-polyacrylamide gel and transferred to PVDF membranes (Bio-Rad, Lab Inc. USA). The membranes were blocked and then incubated with primary antibodies (1:100): anti-phospho-STAT-1 (p- STAT-1; SC-136229), anti-STAT (SC-464), anti-p65 (SC-136548), anti-p38 (SC-7973), and anti-β actin (SC-47778) at 4 °C overnight and then with secondary antibodies labeled with horseradish peroxidase-conjugated mouse anti-human IgG (Sigma) at room temperature 2 hours. The signals were detected using enhanced chemiluminescence reagents (Thermo Scientific, USA). The relative expression was performed by normalizing the intensity of the actin band and adjusting the intensity of the expression in M1-macrophage (control) to 1 unit; subsequently, the intensities of the bands of the treated samples were obtained and compared based on M1-macrophage control and analyzed with the ImageJ 1.53e software (NIH, USA)

### 2.5 LAIR-1 Binding Assays

Binding assays were performed by incubating various concentrations of recombinant human LAIR-1 (R&D #2664-LR-050) overnight at 4°C in 96 micro-wells plates coated with 5 μg/ml native porcine type I collagen or PTIC and blocked with 5% fat-free milk-PBS. Excess protein was removed by washing with PBS containing 0.05% Tween 20. Subsequently, a 1:500 dilution mouse anti-LAIR-1 human (HycultBiotech # HM2364- 100UG) was added overnight at 4°C. Then, it was incubated with anti-IgG mouse labeling HP. The plates were developed with P-NPF. Optical density (OD) was quantified with a microplate reader at 450 nm. Ovalbumin was used as a control and did not bind LAIR-1.^23^ The affinity constant (KA) of the receptor to ligand was used in the model described by Irving Langmuir,^24^ where affinity is inversely proportional to the potency of the ligand (1/Kd).

### 2.6 Patients and Serum samples

Forty sera were obtained from a previous study. It was a single-center, double-blind, placebo-controlled, randomized clinical trial that compared PTIC to placebo in adult outpatients with confirmed COVID-19. The study was approved by the institutional review board of the Instituto Nacional de Ciencias Médicas y Nutrición Salvador Zubirán (INCMNSZ, reference no. IRE 3412-20-21-1) and was conducted in compliance with the Declaration of Helsinki,^25^ the Good Clinical Practice guidelines, and local regulatory requirements. All participants provided written informed consent before being randomly allocated to PTIC or a matching placebo. This study is registered with the ClinicalTrials.gov identifier NCT04517162.

The diagnosis was based on suggestive symptoms (fever, headache, cough, or dyspnea, plus at least another symptom such as malaise, myalgias, arthralgias, rhinorrhea, throat pain, conjunctivitis, vomiting, or diarrhea) and a positive real-time reverse-transcription polymerase chain reaction result for SARS-CoV-2. Subjects that fulfilled the above criteria and whose symptoms started within the previous seven days were included. Exclusion criteria were: hypersensitivity to PTIC or any of its excipients; COVID-19 patients that required hospitalization; all pregnant or breast-feeding women; patients with chronic kidney disease (estimated glomerular filtration rate less than 60 for more than three months or need for hemodialysis or hemofiltration); decompensated liver cirrhosis; congestive heart failure (New York Heart Association class III or IV); and patients with cerebrovascular disease, autoimmune disease, cancer, multiorgan failure, or immunocompromise (solid organ transplant recipient or donor, bone marrow transplant recipient, AIDS, or treatment with biologic agents or corticosteroids). Patients were evaluated by staff at the study site (S.M-F, A.P-R, D.A-Ll, H.O-P, E.O-H, E.R-C.) on days 8, 15, and 97 (1, 7, and 90 days after the last dose of PTIC or placebo, respectively), and patients were encouraged to complete questionnaires daily.

Individuals were asked to provide personal information (date of birth, type of job, educational level, previous contact with infected individuals), pre-existing conditions (systemic hypertension, diabetes mellitus, cardiovascular disease, cerebrovascular disease, hypertriglyceridemia, dyslipidemia), and symptoms. Personal data, exposure history, clinical presentation, chest computed (CT) tomography, laboratory tests, previous treatment, and outcome data were collected prospectively and from inpatient medical records. Laboratory data collected from each patient from the study at baseline, 8 (day one post-treatment), 15 (day eight post-treatment), and 97 days (day 90 post-treatment) included complete blood count, coagulation profile, serum biochemical tests (including renal and liver function tests, electrolytes, lactate dehydrogenase, D dimer, and creatine kinase), serum ferritin, C-reactive protein (CRP) and procalcitonin, basic spirometry, and chest CT scans were done in all patients at baseline, post-treatment and 3months follow-up.

Patients were randomized in a 1:1 fashion to PTIC or placebo. All outcome assessors, investigators, and research staff who interacted with participants were blinded to participant treatment assignment.

Participants received an intramuscular dose of either PTIC (1.5 ml, equivalent to 12.5 mg of collagen) every 12 h for three days and then every 24 h for four days or a placebo. Only acetaminophen or acetylsalicylic acid was allowed as concomitant therapy. Monitoring of compliance was evaluated by counting empty vials returned on subsequent visits.

### 2.7 Serum cytokines

Serum samples were collected from patients treated with PTIC and placebo at 0-, 1-, 8-, and 90 days post-treatment, according to our results in the previous work.^15^ Cytokines were evaluated using the kit Bio-Plex Precision Pro Human Cytokine panel 48-Plex (Bio-Rad, Lab Inc. USA). The samples were processed according to the manufacturer’s manual and read using Bio-Plex 200 System with Bio-Plex Manager 6.1 Software (Bio-Rad, Lab Inc. USA).

### 2.8 Peripheral blood mononuclear cells isolation and flow cytometry

A venous blood sample (10 ml) from each patient and 20 healthy subjects from the blood bank were drawn to perform flow cytometry analysis. Peripheral blood mononuclear cells (PBMCs) were obtained by gradient centrifugation on Lymphoprep (Axis-Shield PoC AS, Oslo, Norway). The cell pellet was resuspended in 1 ml RPMI at 1-2 X 10^6^ cells/ml. PBMCs were incubated with 5 μL of Human TruStain FcXTM (BioLegend Inc.) per million cells in 100 μl PBS for 10 minutes, and then they were labelled with 2 µL of anti-human: (a) CD4 FITC, CD183 PeCy5 and CD192 APC; (b) CD86 FITC, CD11c PeCy5, and CD3 APC; or (c) CD11b FITC, CD16 PeCy5, and CD163 APC antibodies in separated tubes during 20 min at 37°C in the dark. Cells were permeabilized with 200 µL of cytofix/cytoperm solution (BD Biosciences) at 4°C for 30 min. Intracellular staining was performed with an anti-human: (a) IFN-γ PE; (b) IP-10 PE; or (c) IDO PE-labeled mouse monoclonal antibodies for 30 min at 4°C in the dark. An electronic gate was made for live cells (FCSA vs. FCSH), then a) CD4^+^/CD183^+^/CD192^+^; (b) CD86^+^/CD11c^+^/CD3^-^; or (c) CD11b^+^/CD16^+^/CD163^+^ cells.

Results are expressed as the relative percentage of IFN-γ^+^, IP-10^+^, and IDO^+^-expressing cells in each gate.

### 2.9 Chest CT

A semi-quantitative scoring system was used to estimate pulmonary involvement based on the affected pulmonary area.^26, 27^

### 2.10 Basic spirometry

Before the forced expiration, tidal (normal) breaths were taken first, then a deep breath while still using the mouthpiece, followed by a further quick, full inspiration. For FVC and FEV1, the patient took a deep breath in as long as possible, blew out as hard and fast as possible, and kept going until no air was left. PEF was obtained from the FEV1 and FVC maneuvers. For VC, the patient takes a deep breath in, as large as possible, and blows steadily for as long as possible until there is no air left. Nose clips were essential for VC as air can leak out due to the low flow. The IVC maneuver was performed at the end of FVC/VC by taking a deep, fast breath after breathing.

### 2.11 Statistical analysis

A descriptive analysis was done. Continuous variables were expressed by means and standard deviations (normal distribution) or medians, and categorical variables were summarized using proportions. The student’s t-test or the Wilcoxon rank sum test was used for the inferential analysis of continuous variables.

## 3 Results

### 3.1 Differentiation of THP-1 to macrophage-like cells and polarization to M1 macrophages

THP-1 cells were stimulated with PMA, inducing a macrophage-like phenotype. Morphological changes were observed at 72 hours. THP-1 cells changed from cells in suspension (Figure 1A) to adherent cells (Figure 1B). For polarization to M1 macrophage, MLCs were stimulated with IFN-γ and LPS. Polarization was verified by the expression of CD36, CD86, and IL-1β (Figure 1E) compared to the unstimulated MLCs (Figure 1C). Therefore, the results establish that the MLCs were differentiated into M1 macrophages.

**FIGURE 1.**
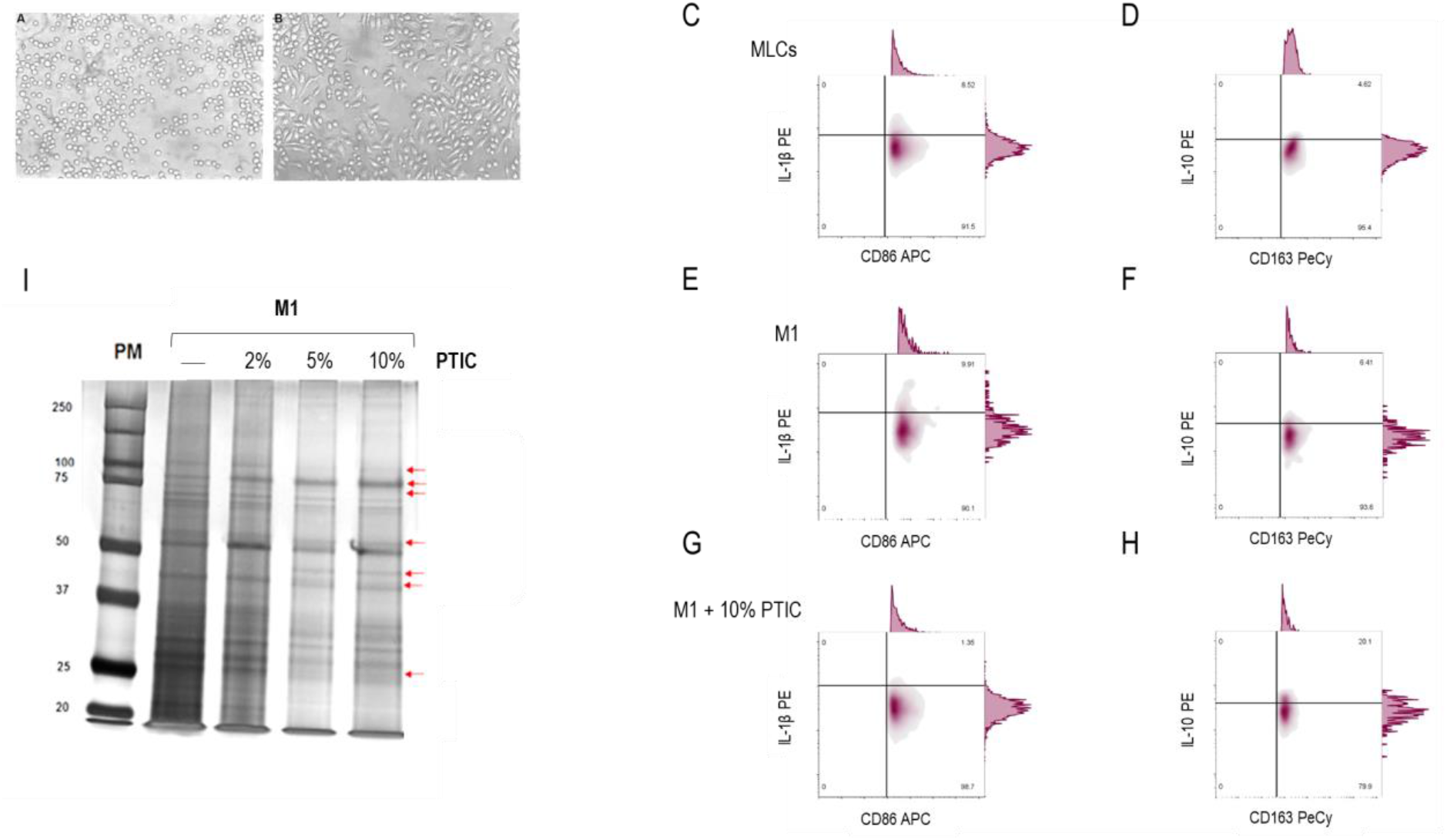
Effect of polymerized type I collagen (PTIC) on M1-macrophages. (A) THP-1 cells. (B) THP-1 cells stimulated with 100 nM PMA for 72 h (monocyte-like cells, MLCs). Characterization of M1 macrophages (CD16^+^/CD36^+^/CD86^+^/IL-1β^+^) in C) MLCs, (E) M1-macrophages (MLCs incubated with 20 ng/ml of IFN-γ and 1μg/ml of LPS for 24 h), and G) M1-macrophages treated with 10% of PTIC. Characterization of M2 macrophages (CD14^+^/CD16^hi^/CD163^+^/IL-10^+^) in (D) MLCs, (F) M1-macrophages, and (H) M1-macrophages treated with 10% of PTIC. (I) Protein expression curves at different concentrations of PTIC (2%, 5%, and 10%). Arrows depict the electrophoretic shifts.

### 3.2 Effect of polymerized type I collagen on M1-macrophages

To determine the effect of PTIC on M1 macrophages and if it was dose-dependent, M1 cells were cultured with 2, 5, and 10% PTIC in RPMI. The banding pattern was compared by electrophoretic shift. We found that by increasing the percentage of PTIC, the bands of 100 kDa, 75 kDa, 55 kDa, 25 kDa, and 22 kDa were decreased. On the other hand, we found an increase in the 40 kDa and 78 kDa bands (Figure 1I). Based on these observations, we decided to carry out all the assays with PTIC at 10%.

The addition of 10% PTIC to the cultures of M1 macrophages induced a decrease in the expression of CD36, CD86, and IL-1β (Figure 1G) and an increase of CD14, CD16, CD163, and IL-10, favoring the M2 macrophage phenotype (CD14^+^/CD16^hi^/CD163^+^/IL-10^+^- producing cells, Figure 1H), in contrast to PTIC untreated cultures (Figure 1D and F).

### 3.3 Polymerized type I collagen binds to LAIR-1 with a similar affinity as native type I collagen

Type I collagen is the natural ligand for LAIR-1. To analyze whether PTIC is a ligand for LAIR-1, a binding assay in which plates coated with native type I collagen or PTIC were used and incubated with human anti-LAIR-1 antibody. The LAIR-1 affinity for PTIC was similar to native collagen (Figure 2A), with KA to PTIC ∼1.01 x 10^-8^ M and native collagen KA ∼1.16 x 10^-8^ M. Ovalbumin was employed as the negative control. These results suggest that PTIC is a ligand for LAIR.

**FIGURE 2.**
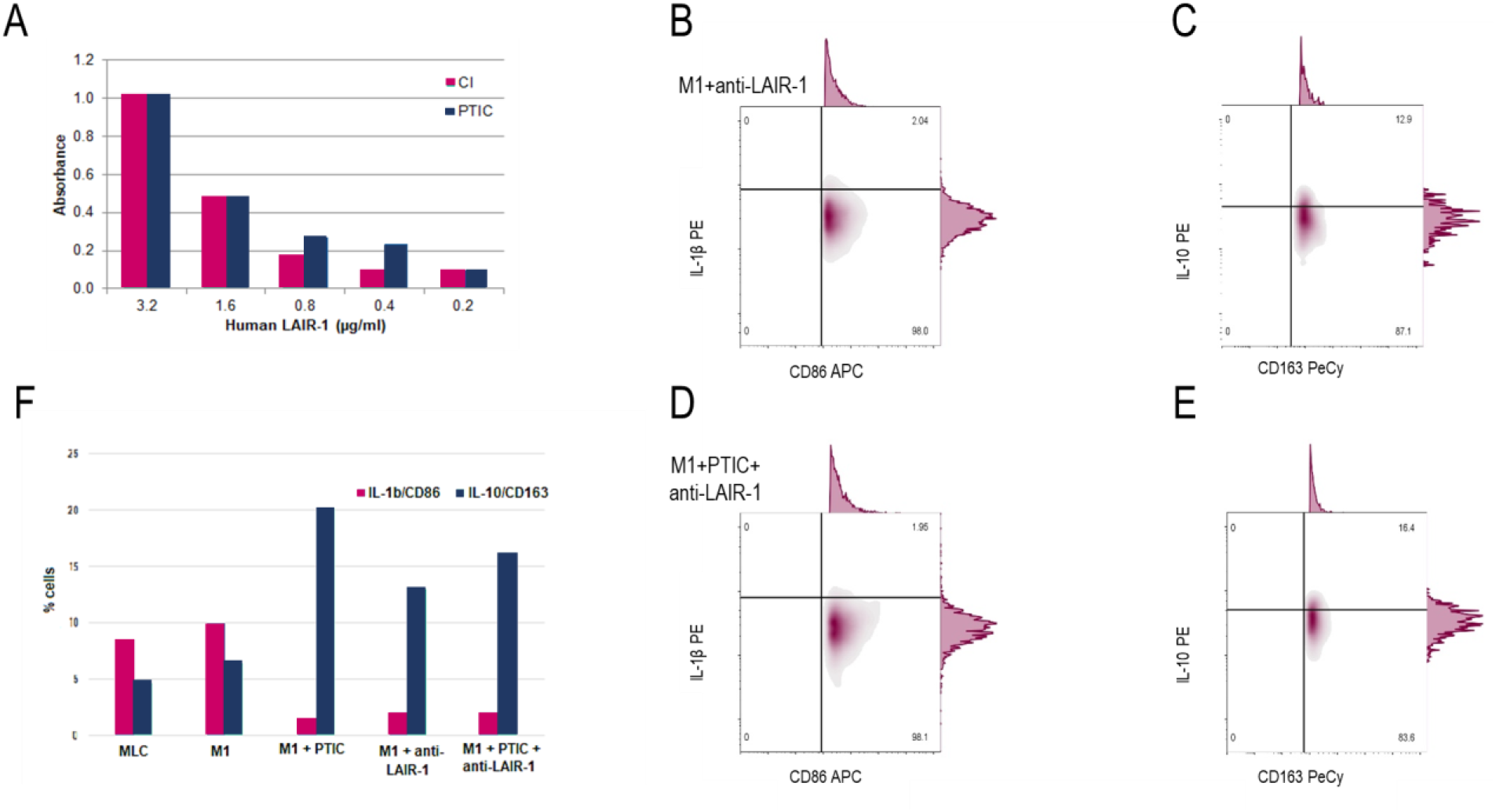
Binding of PTIC to LAIR-1 and its effect on M1-macrophages. (A) Binding assay of PTIC and native porcine type I collagen (CI) to LAIR-1. M1-macrophages treated with (B) anti-LAIR-1 antibody (1:100) for 24 hs or (D) anti-LAIR-1 antibody (1:100) and 10% PTIC for 24 h. Expression of CD14, CD16, CD163, IL-10 in cultures of M1-macrophage treated with (C) anti-LAIR-1 antibody or (E) anti-LAIR-1 antibody and 10% PTIC for 24 h. (F) M1 (CD16^+^/CD36^+^/CD86^+^/IL-1β^+^) and M2 (CD14^+^/CD16^hi^/CD163^+^/IL-10^+^) cell percentage.

#### Activation of LAIR-1 and its effect on M1-macrophages

The addition of anti-LAIR-1 antibody to M1-macrophages cultures decreased expression of M1 markers (CD36, CD86 and IL-1β; Figure 2B and F), and increased M2 markers (CD14, CD16, CD163, and IL-10; Figure 2C and F). A similar phenotype was observed in M1 cultures with anti-LAIR-1 antibody and 10% PTIC (Figure 2D, E, and F). This suggests that PTIC promotes M2 macrophages more than the M1 phenotype.

### 3.4 Signaling pathways regulated by PTIC on M1-macrophages

To determine the signaling pathway regulated by PTIC binding to the LAIR-1 receptor, the lysates from M1 macrophages, M1 cells treated with 2 or 10% PTIC, M1cells activated with IFN-γ, or LPS and THP-1 cells were obtained. They were analyzed by western blot to identify the transcription factors NF-κB (p65), p38, and STAT-1. Adding 2 or 10% PTIC to M1 macrophage cultures did not alter NF-κB (p65) or p38 expression (*data not shown*). This suggests that none of these pathways participates in the LAIR-1 signaling pathway by PTIC. Nevertheless, in cultures of M1 macrophages treated with PTIC, a dose-dependent decrease in STAT-1 phosphorylation was determined. This suggests that PTIC could exert a negative regulatory effect on the inflammation of M1-macrophages, inhibiting the stimulus signaling by IFN-γ (Figures 3A and B).

**FIGURE 3.**
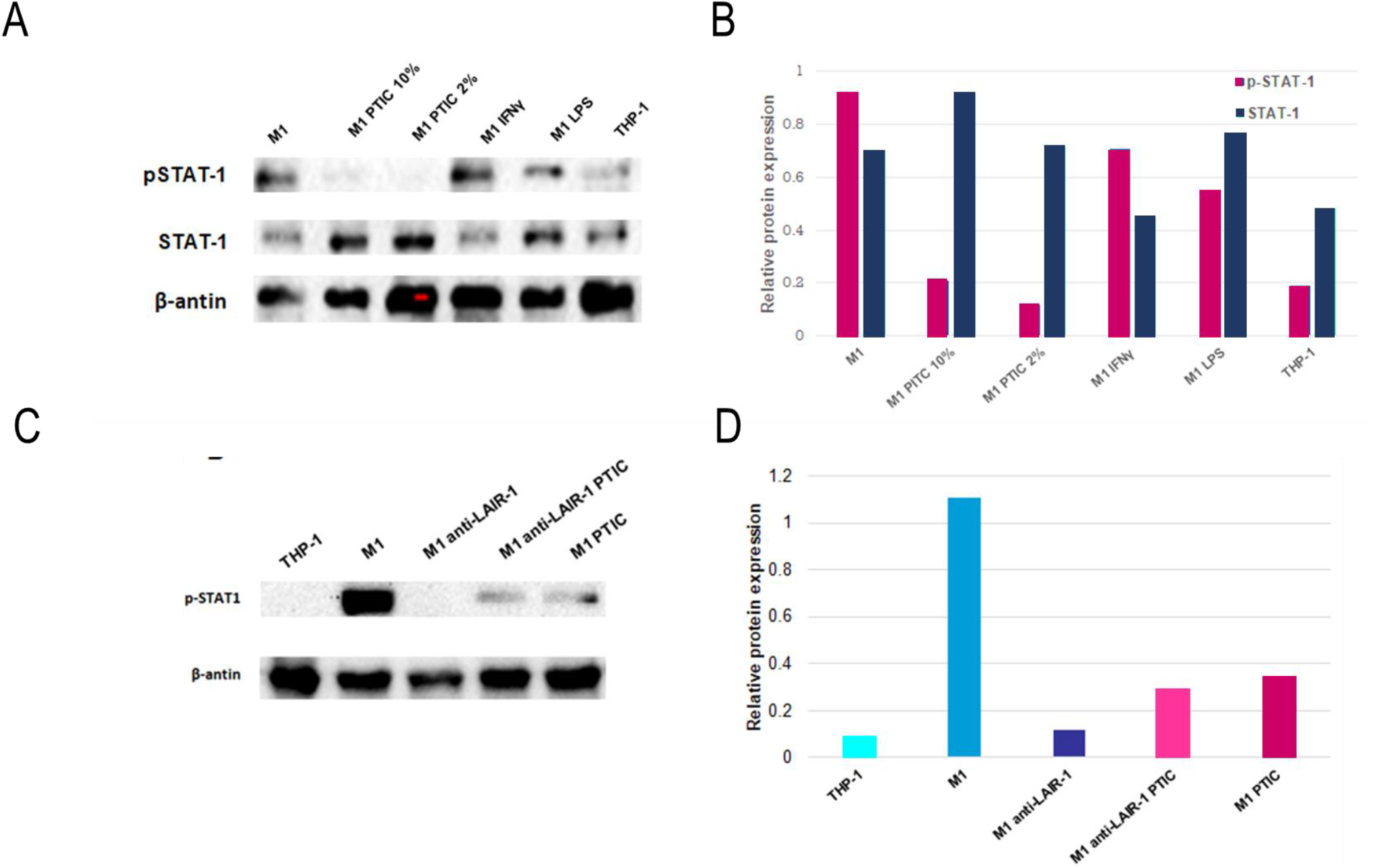
Binding of PTIC to LAIR-1 induces downregulation of STAT-1 phosphorylation. (A) Western blotting relative expression of STAT-1 and p-STAT1. (B) Relative expression of STAT-1 and pSTAT-1. Both were normalized for actin and were adjusted to 1 unit. (C) Effect of STAT-1 phosphorylation by activating LAIR-1 with anti-LAIR-1 antibody in M1-macrophages. (D) Relative expression of phosphorylated STAT-1. Results were normalized for actin and were adjusted to 1 unit.

Adding an anti-LAIR-1 antibody to M1macrophages cultures activated the receptor, reducing STAT-1 phosphorylation more efficiently than PTIC or the combination of anti-LAIR-1 and PTIC (Figure 3C and D).

### 3.5 Baseline description of the study population

Forty adult non-hospitalized patients with COVID-19 (mild to moderate disease) were included in the study. The mean (±SD) age of the patients was 49.6±13.8 years. Twenty patients (50%) were male. According to the Guangzhou score to predict the occurrence of critical illness, the mean score was 93.2±24.4 (medium risk). The mean (±SD) oxygen saturation of study participants was 91.8±2.9. Sixteen patients (40%) had an oxygen saturation of 91% or lowered while breathing ambient air (6 of the PTIC group and 10 of the placebo group). Coexisting conditions and symptoms are described in Table 1.

**TABLE 1.**
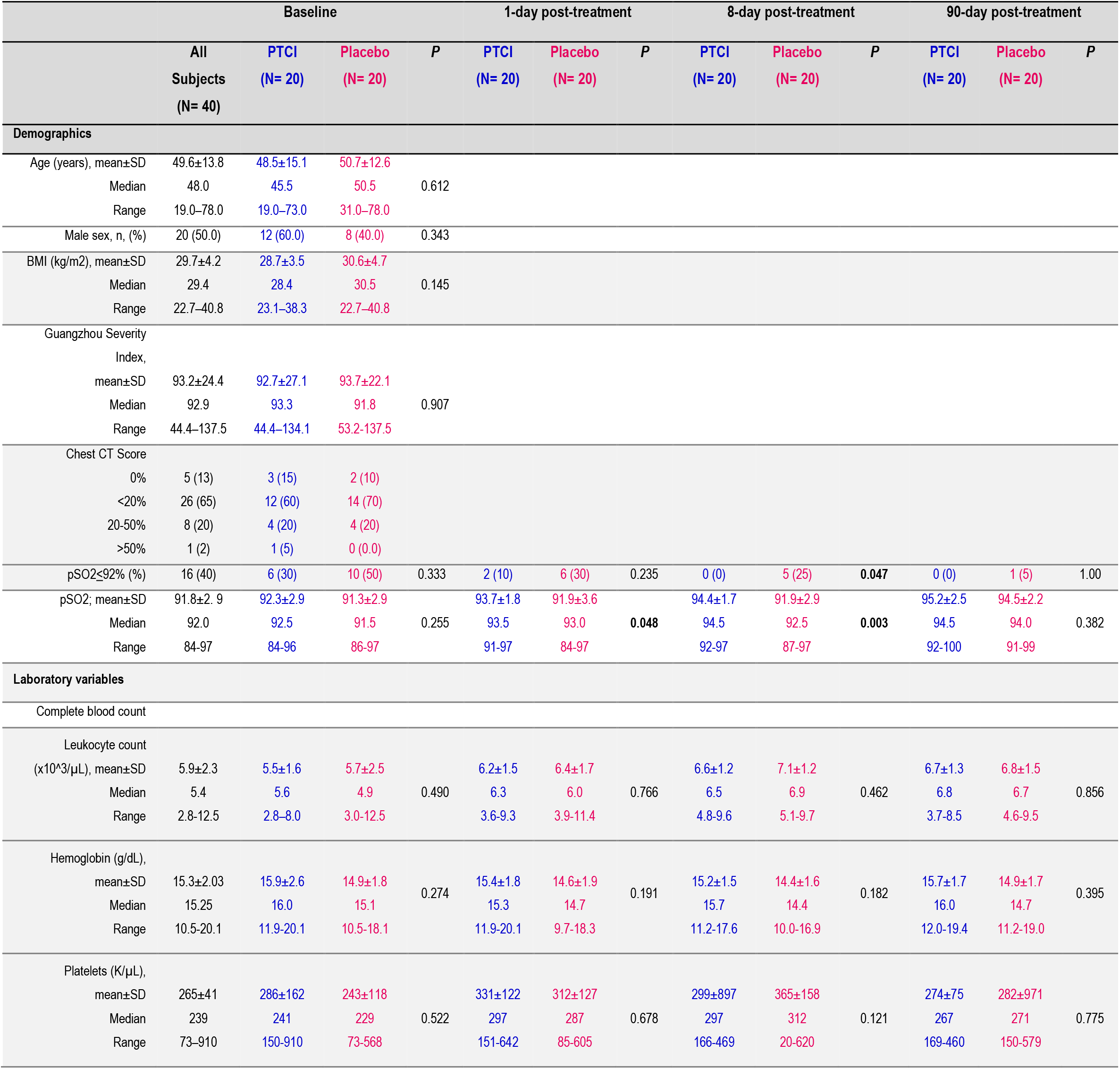
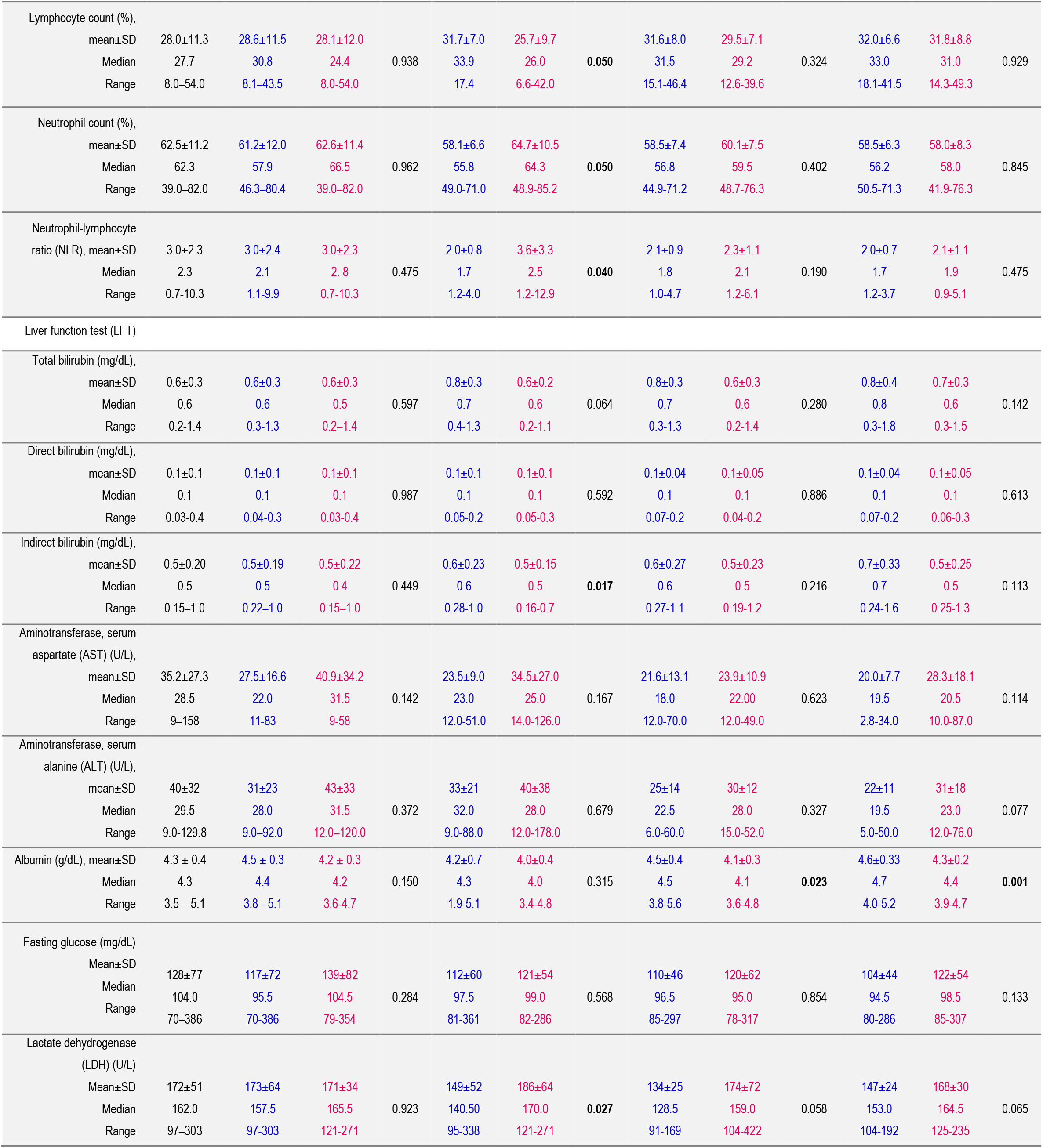
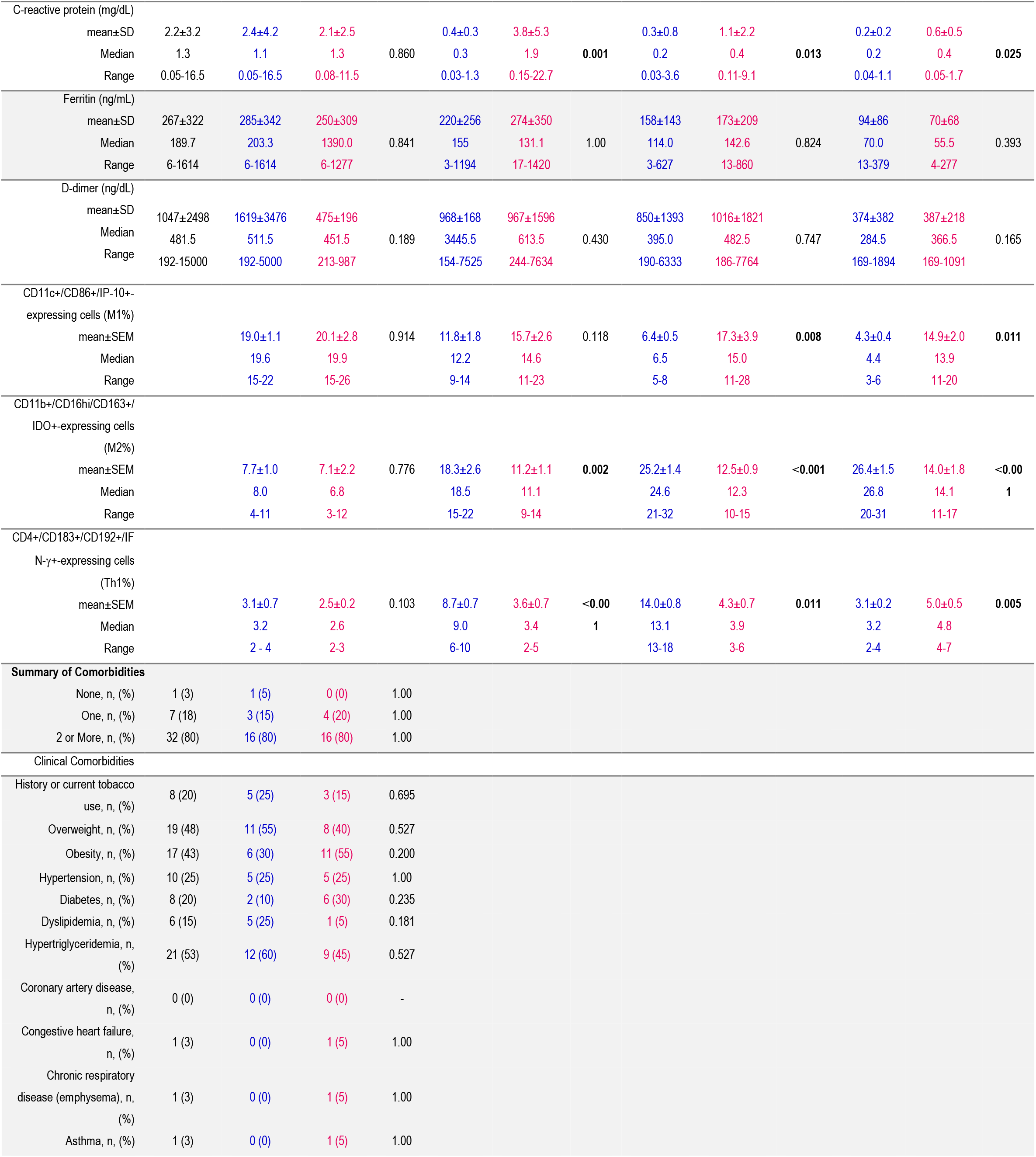
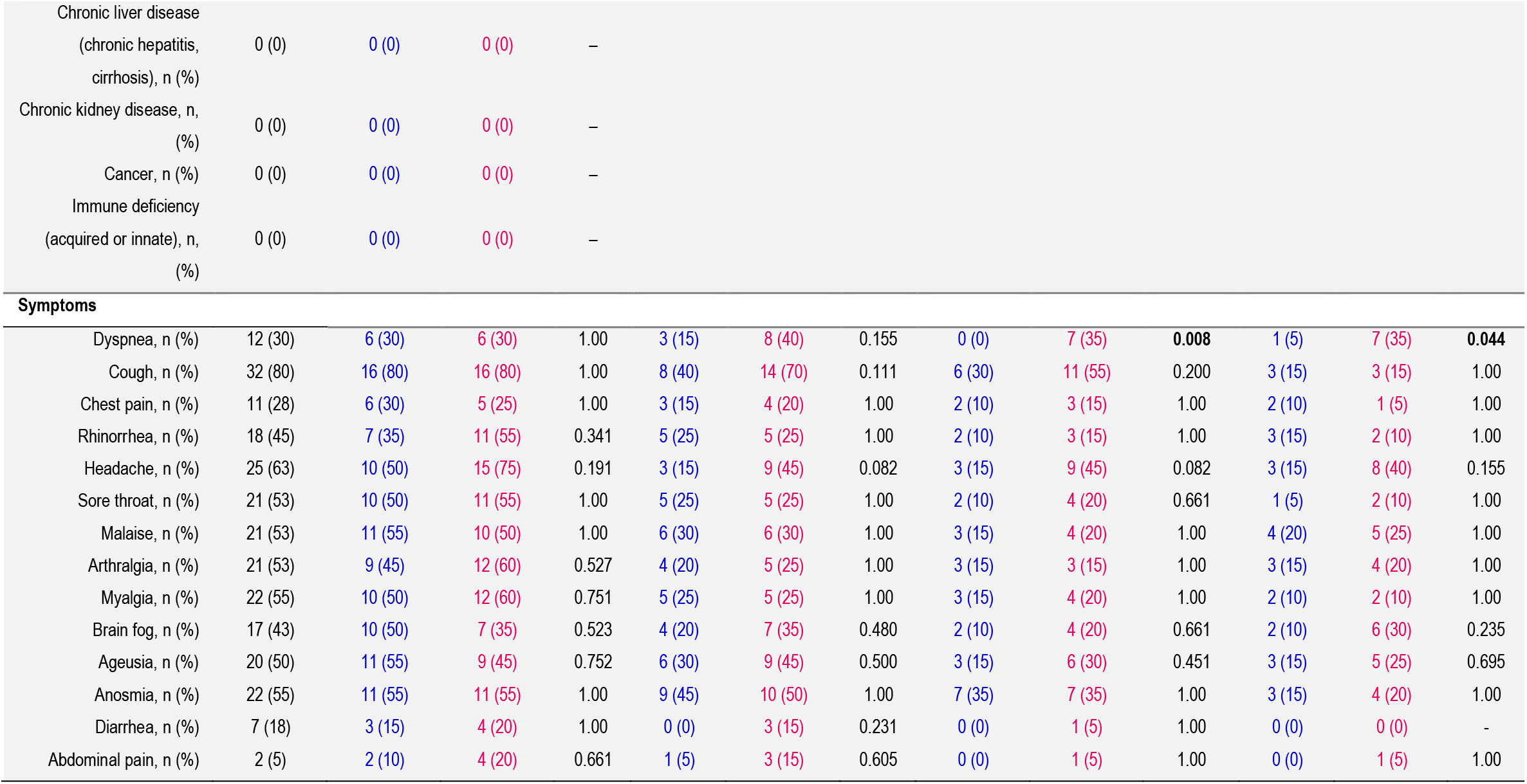
Demographic and clinical characteristics of the trial population.

Regarding radiological abnormalities on chest CT, 35 patients (87%) had lung disease; of these, 26 (65%) had less than 20% lung parenchymal involvement, 8 (20%) had between 20 and 50%, and 1 (2%) had higher than 50% (Table 1).

### 3.6 Concomitant medications

Of 40 patients at baseline, 28 (70%) were being treated with acetaminophen, 13 (33%) with acetylsalicylic acid, 3 (8%) with antivirals (oseltamivir), and 16 (40%) with antibiotics (azithromycin, ceftriaxone, penicillin, clarithromycin, and levofloxacin). The use of acetaminophen (35% *vs.* 35%), acetylsalicylic acid (20% *vs.* 13%), antivirals (5% *vs.* 3%), and antibiotics (22% *vs*. 18%) were similar in the PTIC and placebo groups, respectively. No patients were treated with anticoagulants or steroids.

### 3.7 Percentage of circulating M1 and M2 macrophages in patients with COVID-19 treated with PTIC

During the early phase of the infection, a high percentage of M1 cells was determined, which decreased to statistically significant levels from day 1 to 90 post-treatment in patients treated with PTIC but not in placebo, where no change in levels was observed in this subset regarding baseline (Figure 4B-G; Table 1). Differences in the number of circulating M1 at day 15 and 90 post-treatment between the PTIC-treated group and placebo were determined (Figure 4G; Table 1).

**FIGURE 4.**
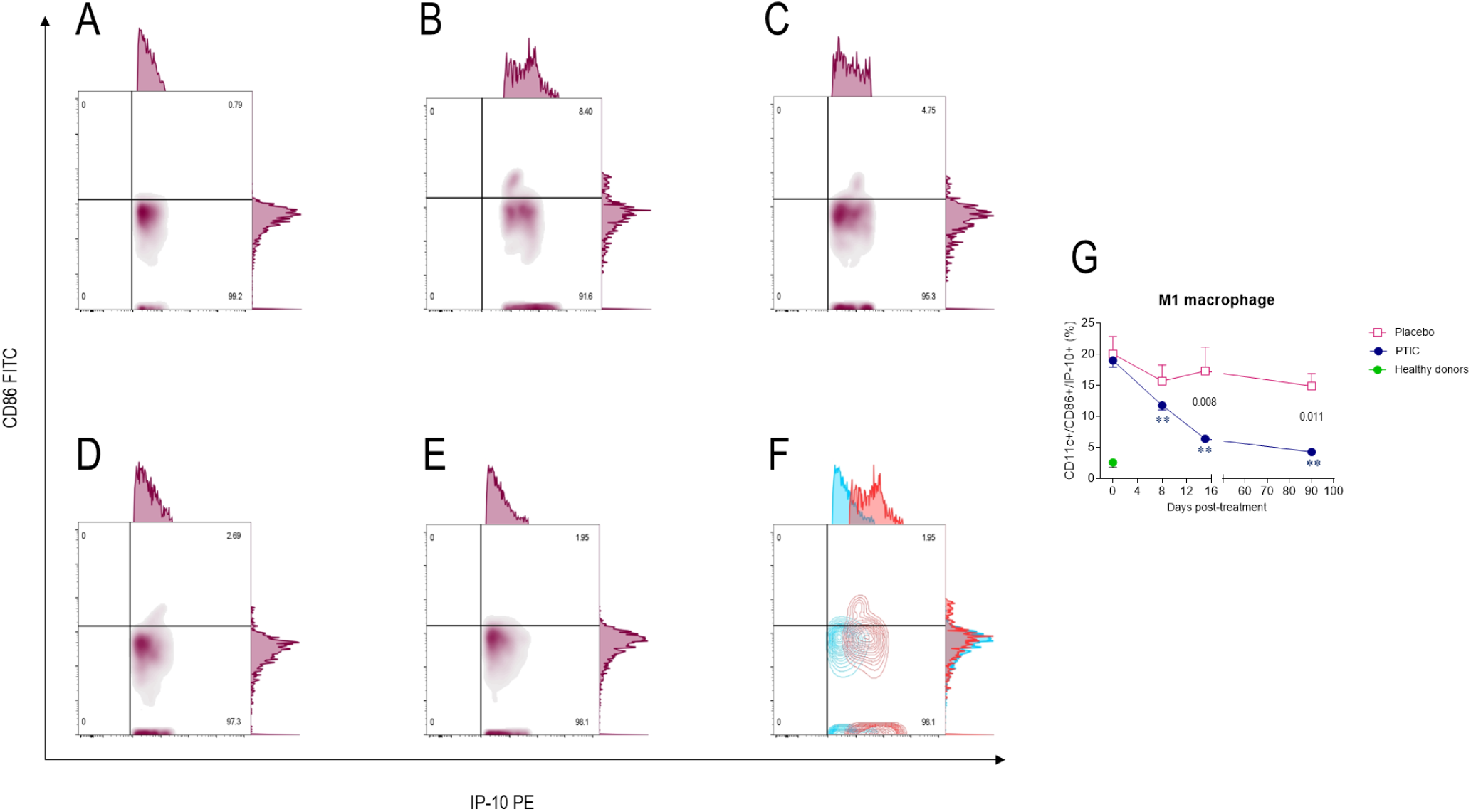
Representative flow plots of circulating M1 subset in SARS-CoV2-infected symptomatic outpatients at baseline 1-, 8-, and 90-days post-treatment with PTIC or placebo. CD86^+^/CD11c^+^/CD3^-^/IP-10^+^- expressing cells in (A) healthy donors; and patients at (B) baseline, (C) 1 day, (D) 8 days, and (E) 90 days post-treatment. (F) Flow plots at baseline (red) and 90 days post-treatment with PTIC (blue). (G) Data are expressed as mean ± SEM. Blue stars show the day the treatment reached a P < 0.05 compared to the PTIC treatment baseline. Pink stars depict the day the therapy reached a *P* < 0.05 compared to the baseline for the placebo. The numbers on the graph represent the statistical significance between the patients treated with PTIC vs. placebo. *P≤0.05 **P≤0.001 depict the statistically significant difference from baseline (blue: PTIC, pink: placebo, green: healthy donors).

In contrast, the percentage of M2 cells increased to statistically significant levels from day 1 to 90 post-treatment in patients treated with PTIC vs. placebo (Figure 5B-G; Table 1). Differences were found between the treatments in the number of circulating M2 at day 1 and 90 post-treatment (Figure 5G; Table 1).

**FIGURE 5.**
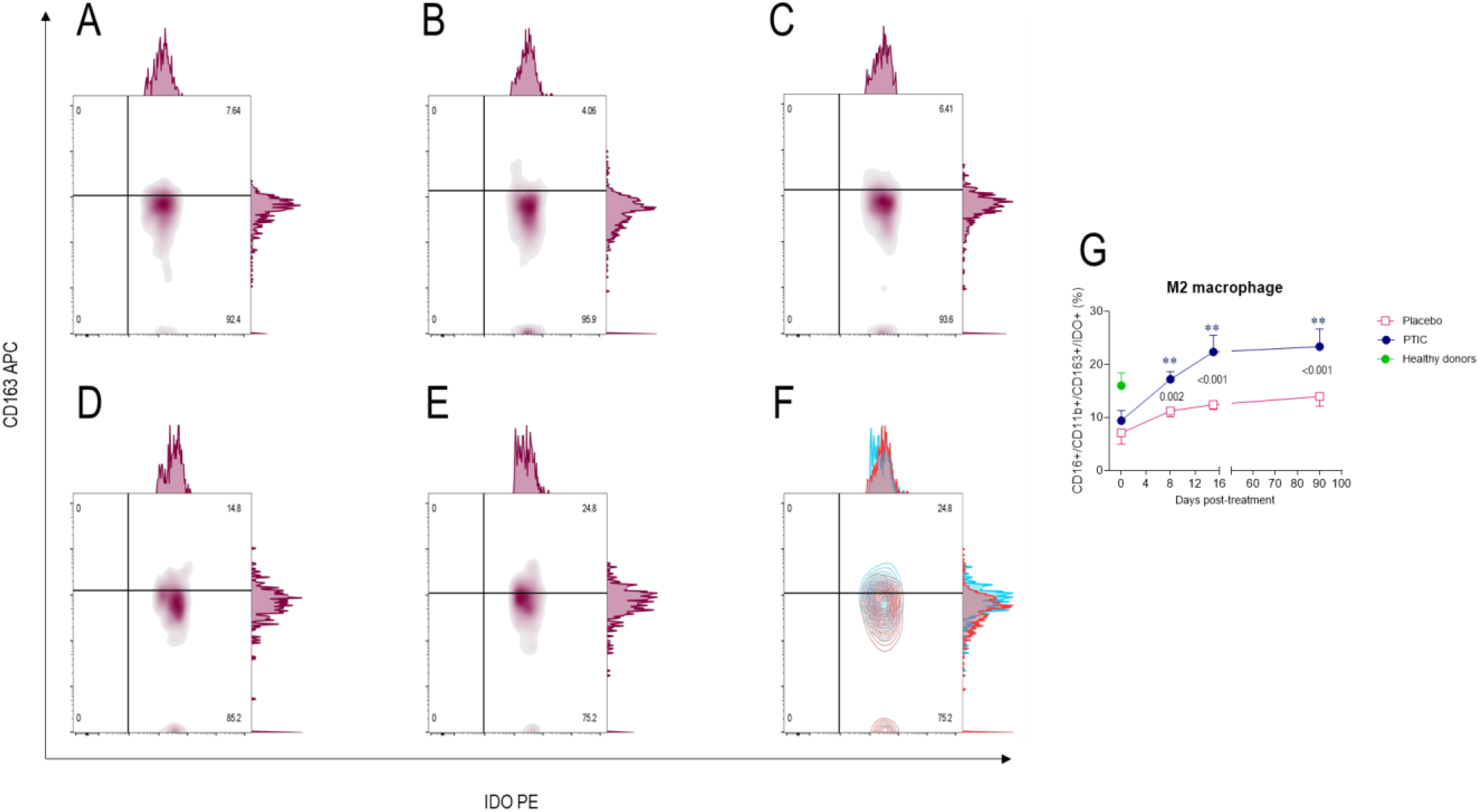
Representative flow plots of circulating M2 subset in SARS-CoV2-infected symptomatic outpatients at baseline 1-, 8-, and 90-days post-treatment with PTIC or placebo. CD11b^+^/CD16^+^/CD163^+^/IDO^+^-expressing cells in (A) healthy donors; and patients at (B) baseline, (C) 1 day, (D) 8 days, and (E) 90 days post-treatment. (F) Flow plots at baseline (red) and 90 days post-treatment with PTIC (blue). G) Data are expressed as mean ± SEM. Blue stars show the day the treatment reached a P < 0.05 compared to the PTIC treatment baseline. Pink stars depict the day the treatment reached a *P* < 0.05 compared to the baseline for the placebo. The numbers on the graph represent the statistical significance between the patients treated with PTIC vs. placebo. *P≤0.05 **P≤0.001 depict the statistically significant difference from baseline (blue: PTIC, pink: placebo, green: healthy donors).

### 3.8 Percentage of circulating Th1 cells in patients with COVID-19 treated with PTIC

IFN-γ-producing effector CD4 T cells increased from day 1 after the last administration of PTIC and decreased to levels similar to those of the control group by day 90 post-treatment. In contrast, Th1 cells remained low in those patients treated with the placebo (Figure 6A-G; Table 1). Differences were determined between the treatments in the number of circulating Th1 at day 1 and 15 post-treatment (Figure 6G; Table 1).

**FIGURE 6.**
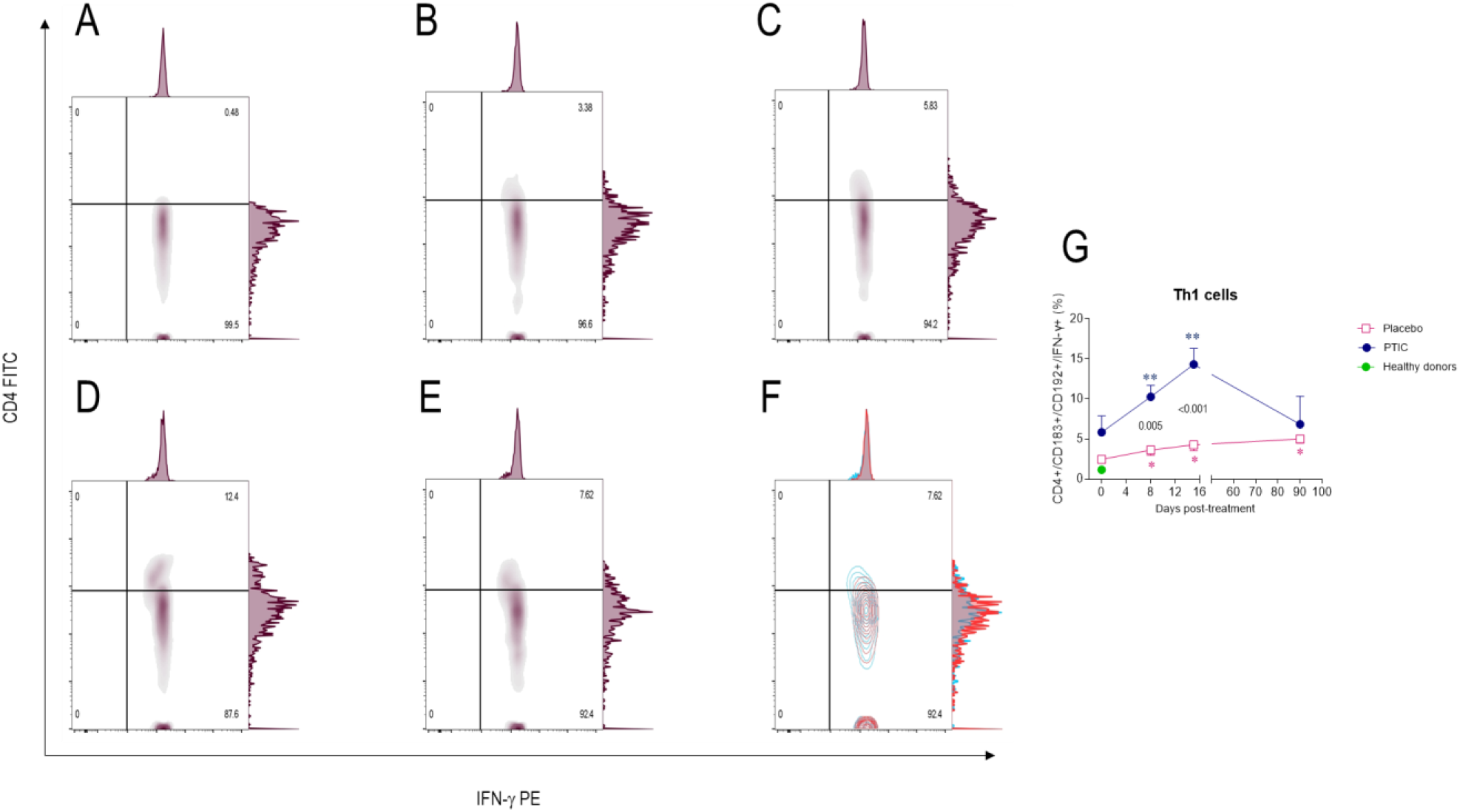
Representative flow plots of circulating Th1 subset in SARS-CoV2-infected symptomatic outpatients at baseline 1-, 8-, and 90-days post-treatment with PTIC or placebo. CD4^+^/CD183^+^/CD192^+^/IFN-γ^+^-expressing cells in (A) healthy donors; and patients at (B) baseline, (C) 1 day, (D) 8 days, and (E) 90 days post-treatment. (F) Flow plots at baseline (red) and 90 days post-treatment with PTIC (blue). (G) Data are expressed as mean ± SEM. Blue stars show the day the treatment reached a P < 0.05 compared to the PTIC treatment baseline. Pink stars depict the day the treatment reached a *P* < 0.05 compared to the baseline for the placebo. The numbers on the graph represent the statistical significance between the patients treated with PTIC vs. placebo. *P≤0.05 **P≤0.001 depict the statistically significant difference from baseline (blue: PTIC, pink: placebo, green: healthy donors).

### 3.9 Serum cytokine and chemokines levels

To determine cytokine levels in patients, we followed up and measured serum cytokine levels in patients treated with PTIC and those who received the placebo on days 0, 1, 8, and 90. We found a significant decrease in IP-10 (*P*<0.001; Figure 7A), IL-8 (*P* <0.001; Figure 7D), M-CSF (*P* =0.021; Figure 7F), HGF (*P* = 0.030; Figure 7G) while there was an increase in stem cell factor (SCF, *P* = 0.005; Figure 7E) and tumor necrosis factor (TNF)- related apoptosis-inducing ligand (TRAIL, *P* = 0.003; Figure 7H) on day 1 of PTIC treatment. On the other hand, only the migration inhibitory factor (MIF, p=0.049; Figure 7B) showed a decrease on day 8, and only Eotaxin decreased on day 90 (*P* = 0.015; Figure 7C).

**FIGURE 7.**
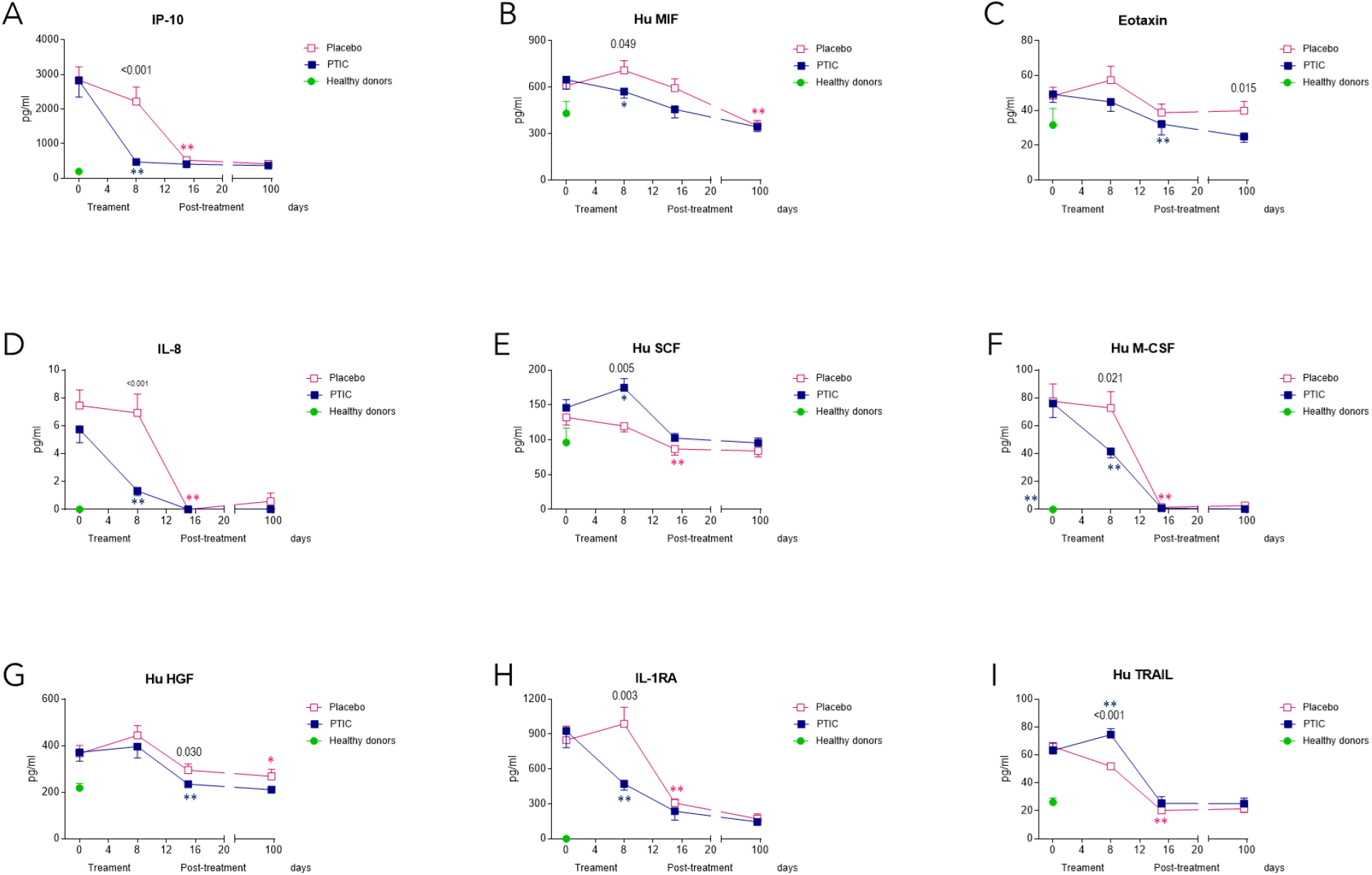
Serum cytokine and chemokine levels of SARS-CoV2-infected symptomatic outpatients at baseline 1-, 8-, and 90-days post-treatment with PTIC or placebo. Data are expressed as mean ± SEM. (A) IP- 10, (B) Hu MIF, (C) Eotaxin, (D) IL-8, (E) Hu SCF, (F) Hu M-CSF, (G) Hu HGF, (H) IL-1Ra, and (I) Hu TRAIL. The numbers on the graph represent the statistical significance between the patients treated with PTIC vs. those treated with a placebo. *P≤0.05 **P≤0.001 depict the statistically significant difference from baseline (blue: PTIC, pink: placebo, green: healthy donors).

### 3.10 Oxygen saturation

On days 1, 8, and 90 post-treatment, the percentage reported by the subjects with oxygen saturation readings ≥92% in the PTIC and placebo groups were 90 vs. 70%, 100 vs. 75% (*P*=0.047), and 100 vs. 95%, respectively (Table 1). The mean oxygen saturation in the PTIC and placebo groups in the time above points were 93.7±1.8 vs. 91.9±3.6 (*P*=0.048), 94.4±1.7 vs. 91.9±2.9 (*P*=0.003), and 95.2±2.5 vs. 94.5±2.2 (*P*=0.382), respectively (Figure 8A, Table 1).

**FIGURE 8.**
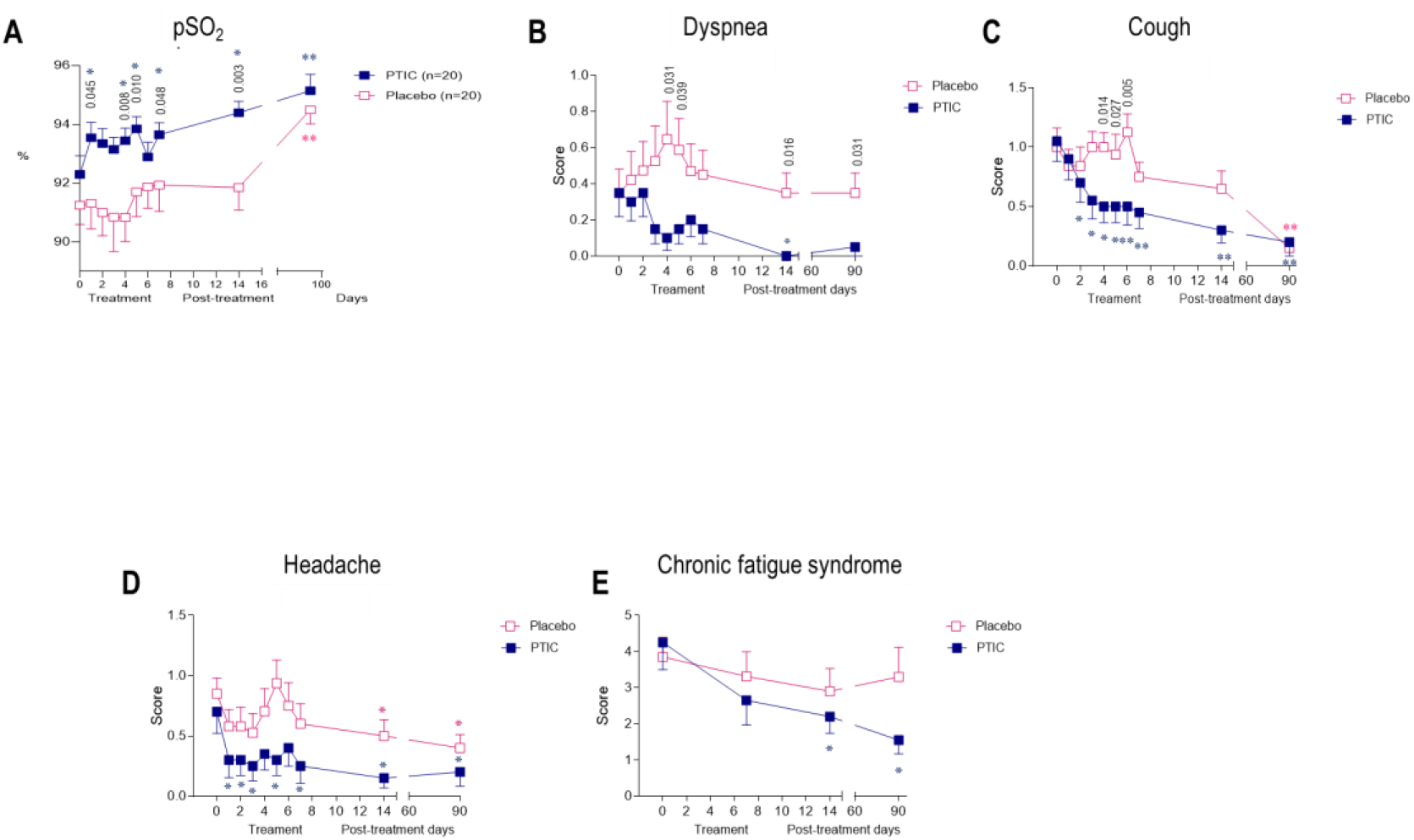
(A) Oxygen saturation of SARS-CoV2-infected symptomatic outpatients at baseline 1-, 8-, and 90-days post-treatment with PTIC or placebo. The numbers on the graph represent the statistical significance between the patients treated with PTIC and those treated with placebo .*P ≤0.05 **P≤0.001 depict the statistically significant difference from baseline (blue: PTIC, pink: placebo). The intensity of symptoms during treatment and follow-up of outpatients with symptomatic COVID-19 treated with PTIC or placebo. (B) Dyspnea, (C) cough, (D) headache, (E) chronic fatigue syndrome evaluated by chalder fatigue questionnaire (The bimodal evaluation produces a score from 0 to 11. A score greater than or equal to 4 qualifies as a “case”). The intensity of the symptom was evaluated on a 4-point rating scale (0 = without symptom, 1 = mild, 2 = moderate, 3 = severe). Blue lines represent the group of patients under polymerized type I collagen treatment. Red lines represent the group of patients under placebo treatment. Results depict mean ± standard error of the mean. Blue stars show the day the treatment reached a P < 0.05 compared to the PTIC treatment baseline. Pink stars depict the day the treatment reached a *P* < 0.05 compared to the baseline for the placebo.

### 3.11 Chest imaging

Imaging of subjects initially revealed characteristic patchy infiltration, progressing to extensive ground-glass opacities that often presented bilaterally. Abnormalities on chest CT scans were detected among 82% of the patients in this study, and no differences between groups were detected (Table 1). At 90-day post-treatment, 2 (10%) patients in the PTIC group and 3 (15%) patients in the placebo group had cicatricial changes.

Furthermore, 8 (40%) patients in the PTIC group and 10 (50%) patients in the placebo group had pneumonitis. No patient had pneumonia. In contrast, all patients treated with PTIC (11) had normal spirometry.

### 3.12 Basic spirometry

At 90 days post-treatment, 3 of 13 (23%) patients in the placebo group had a mild restrictive pattern.

### 3.13 Symptoms

The symptom improvement of the patients was registered daily and compared with the baseline. Significant improvements in the intensity of dyspnea, cough, headache, and chronic fatigue syndrome were noticed during treatment and follow-up in PTIC subjects (Figure 8B-E).

### 3.14 Adverse events

In this study, no serious adverse events were detected. PTIC was safe and well tolerated.

In the PTIC group, the following were observed on day one after treatment: 13 patients had pain in the injection site lasting 15-20 minutes, and one patient had an urticarial rash at the injection site on day 6. On days 8 and 90 after treatment, no adverse events were reported.

In the placebo group, the following were observed on day one after treatment: 15 patients had pain in the injection site lasting 15-20 minutes, and one patient had abdominal pain. On days 8 and 90, 1 patient had tachycardia.

### 3.15 Laboratory assays

No differences in laboratory results were found among the PTIC and placebo groups at baseline (Table 1).

On days 1, 8, and 90 post-treatment with PTIC, serum levels of high sensitivity CRP (hs- CRP) decreased compared with placebo (*P*≤0.001, *P*=0.013 and *P*=0.025, Table 1).

On day 1 post-treatment with PTIC, serum levels of lactate dehydrogenase decreased compared with placebo (*P*=0.027, Table 1).

On days 8 and 90 post-treatment with PTIC, serum levels of albumin increased compared with the placebo (*P*=0.023, and *P*≤0.001, Table 1).

On day 1 post-treatment with PTIC, neutrophil-to-lymphocyte ratio (NLR) decreased compared with placebo (*P*=0.040, Table 1).

## 4 DISCUSSION

The extracellular matrix is a complex and dynamic structure that in mammals is composed of at least 1100 different proteins, recognized as the matrisome. It is classified into collagens, glycosaminoglycans, proteoglycans, and glycoproteins. The collagen family represents 25 to 30% of all body proteins. In vertebrates, more than 40 genes synthesize α chains, which associate in threes to form up to 29 different types of collagen molecules. Its primary function is to create a support structure resistant to the force of mechanical tension for the tissues. Cells adhere to collagen through adhesion molecules such as integrins, selectins, receptor tyrosine kinases, and molecules from the immunoglobulin family. Collagen is characterized by having a composition rich in glycine (−Gli−X−Y−Gli−X−Y), where “X” and “Y” are usually proline and hydroxyproline, respectively. The most frequent is type I, which represents 90% of the total collagen in the organism.^28^

It has been previously reported that type I collagen is a functional ligand for LAIR-1. The interaction depends on the conserved glycine-proline-hydroxyproline (GPO) repeat region of collagen and a conserved arginine residue on LAIR1 (R59). Thus, the engagement of collagen and LAIR-1 directly inhibits immune cell function.^29^ LAIR-1 is expressed in most hematopoietic cells, and its role has been studied on multiple immune cells, mainly lymphocytes and neutrophils. Nonetheless, several functions of LAIR-1 associated with monocytes/macrophages have been reported. LAIR-1 ligands may inhibit the levels of macrophage inflammatory mediators, including tumor necrosis factor (TNF)- α, macrophage inflammatory protein (MIP)-1, MIP-2, RANTES, and macrophage-induced gene (MIG) and, may modulate apoptosis.^30^ LAIR1 is most highly expressed by nonclassical monocytes (M2), followed by classical monocytes (M1) and tissue-resident macrophages.^31^ LAIR-1 can bind to SHP-1 in NKs, and bind to JAK1 and JAK2, regulating the phosphorylation of STAT-1, subsequently resulting in the down-regulation of immune-related genes.^32^

To analyze the possible mechanism of PTIC treatment on macrophages, we evaluated THP-1 cells differentiated to MLC and polarized to M1-macrophages cultured with different concentrations of PTIC. We assessed the activation of the main signaling pathways NF-kB, p38 (*data not shown*), and STAT-1, and we only observed a significant decrease in STAT-1 phosphorylation when cultivating M1 macrophages with PTIC. This down-regulation seems to favor the polarization towards M2-macrophages (increased IL- 10 and CD163), which could contribute to the repair of damaged tissue and decrease inflammation. It has been reported that macrophages can reverse their polarized phenotypes depending on STAT1 phosphorylation. Thus, the inhibition of the STAT1 phosphorylation suppresses the M1-macrophage polarization and promotes the phosphorylation of STAT6 and the M2-macrophage phenotype.^33^ This would contribute to a less inflammatory microenvironment.

IFN-γ is the main cytokine associated with M1 polarization and the main Th1 cell product. Other cells, such as natural killers and macrophages, have been shown to produce the cytokine.^34^ IFN-γ controls specific gene expression involving cytokine receptors (CSF2RB, IL-15 receptor alpha [RA], IL-12RA, and IL-6R), cell activation markers (CD36, CD38, CD69, and CD97), and several cell adhesion molecules (intercellular adhesion molecule 1 [ICAM- 1], integrin alpha L [ITGAL], and mucin 1 (MUC1)^35^ IFN-γ is a cytokine with essential roles in the immune and inflammatory response. Its signaling activates the Janus kinase (JAK)- signal transducer and activator of the transcription 1 (STAT1) pathway to induce the expression of classical interferon-stimulated genes that essential immune effector functions.^36–38^ The increased proinflammatory activity is characterized by INF-γ-mediated polarization of macrophages to an M1-like state.^38^ Viral infection may induce M1 polarization of macrophages, which is generally considered paramount in viral clearance due to the release of proinflammatory cytokines. However, the M1-alveolar strongly takes up, amplifies, and releases SARS-CoV-2, thus spreading the viral infection, while the M2- alveolar conversely degrades the virus and limits its spread.^39^ The finding of an increase in the circulating Th1 cell subpopulation in the first 15 days post-treatment with PTIC in patients suggests a potential role in viral clearance.

The findings are consistent with those observed in outpatients with COVID-19 treated with PTIC, where there was a decreased serum IP-10, an early marker of M1 macrophages. PTIC can inhibit STAT-1 signaling IFN-γ induced in M1-macrophages, resulting in great importance, and it could elucidate a relevant mechanism in treating PTIC in COVID-19 patients.

Multiple studies have demonstrated the presence of biomarkers in COVID-19 that allow classification by severity. This study analyzed samples from outpatients with COVID-19 treated with PTIC or placebo. In the group treated with PTIC, we found a decrease in IP- 10, IL-8, and M-CSF concentration one day after treatment,^15^ indicating that PTIC could regulate macrophage responses. IP-10 and IL-8 are biomarkers associated with severe disease.^26, 40–43^

SARS-CoV-2 infection induces exuberant inflammatory responses and increased secretion of IL-1β, IFN-γ, IP-10, monocyte chemotactic protein 1 (MCP-1), IL-4, and IL-10. IP-10, GM-CSF, macrophage inflammatory protein (MIP)-2, and MIP-1β are early markers of M1- macrophages.^44^ Further, IL-8 plasma levels were elevated in mild and severe COVID-19 patients and increased with the disease progression.^26^ IL-8 is a proinflammatory cytokine that has a role in neutrophil activation and has been identified within the pathogenesis and progression of this disease as a biomarker and prognostic factor in the modulation of the hyperinflammatory response in acute respiratory distress syndrome.^43^ Numerous studies indicate that IL-8 is expressed in various cell types, including neutrophils, fibroblasts, epithelial cells, hepatocytes, alveolar macrophages, and endothelial cells.^42–43^ The macrophage can be a significant player in the so-called cytokine storm and may damage the tissues. It is believed that SARS-CoV-2 induces macrophage-activation syndrome to be lethal.^27^ For this reason, we decided to study the effect of PTIC on the macrophage, mainly the M1-macrophage, which is proinflammatory, since it directs the cytokine release syndrome.

In this study, it has been demonstrated that PTIC treatment helped decrease the levels of IP-10, IL-8, and M-CSF, all of them biomarkers of severe disease, during the first week of treatment and the follow-up. This finding is relevant since Zhao Y. et al. have described that IP-10 levels decline from week 2 and return to normal in week 4 in the group of patients with mild disease. In contrast, in the severe cases, the IP-10 levels remain high at week 2 and start to decline at week 3, and further by week 4.^45^ Therefore, treatment with PTIC reduces the levels of IP-10 by 70% at week 1 in patients with moderate disease, suggesting its regulatory role on cytokine release syndrome.

Intramuscular PTIC was associated with better oxygen saturation values when compared to placebo. Also, PTIC shortened symptom duration. At days 1, 8, and 90 post-treatment with PTIC, a higher mean oxygen saturation value and a higher proportion of patients retaining oxygen saturation values ≥92% were observed. This could be related to a decrease in dyspnea and cough.^15–18, 46^ It should be noted that patients treated with PTIC did not present chronic fatigue syndrome compared to patients treated with the placebo.

Regarding systemic inflammation, at days 1, 8, and 90 post-treatment with PTIC, statistically significant lower levels of hs-CRP were observed. The benefit was evident in the early stage of the infection (7 days after symptom onset). CRP reflects the total systemic burden of inflammation in several disorders. CRP has been shown to upregulate the production of proinflammatory cytokines and adhesion molecules (ICAM-1, VCAM-1 end ELAM-1), and its expression is regulated by a proinflammatory milieu enriched with IL-6.^35^ High levels of CRP are found closely correlated with disease severity.^47^ Moreover, PTIC was safe, well-tolerated, and effective for improving symptoms in outpatients with mild to moderate COVID-19. It did not induce liver damage, hematopoiesis impairment, or blood count alterations.

Summing up, PTIC is an agonist of LAIR-1 and down-regulates STAT-1 phosphorylation (Figure 9). PTIC could be relevant for treating STAT-1-mediated inflammatory diseases, including COVID-19 and long COVID-19.

**FIGURE 9.**
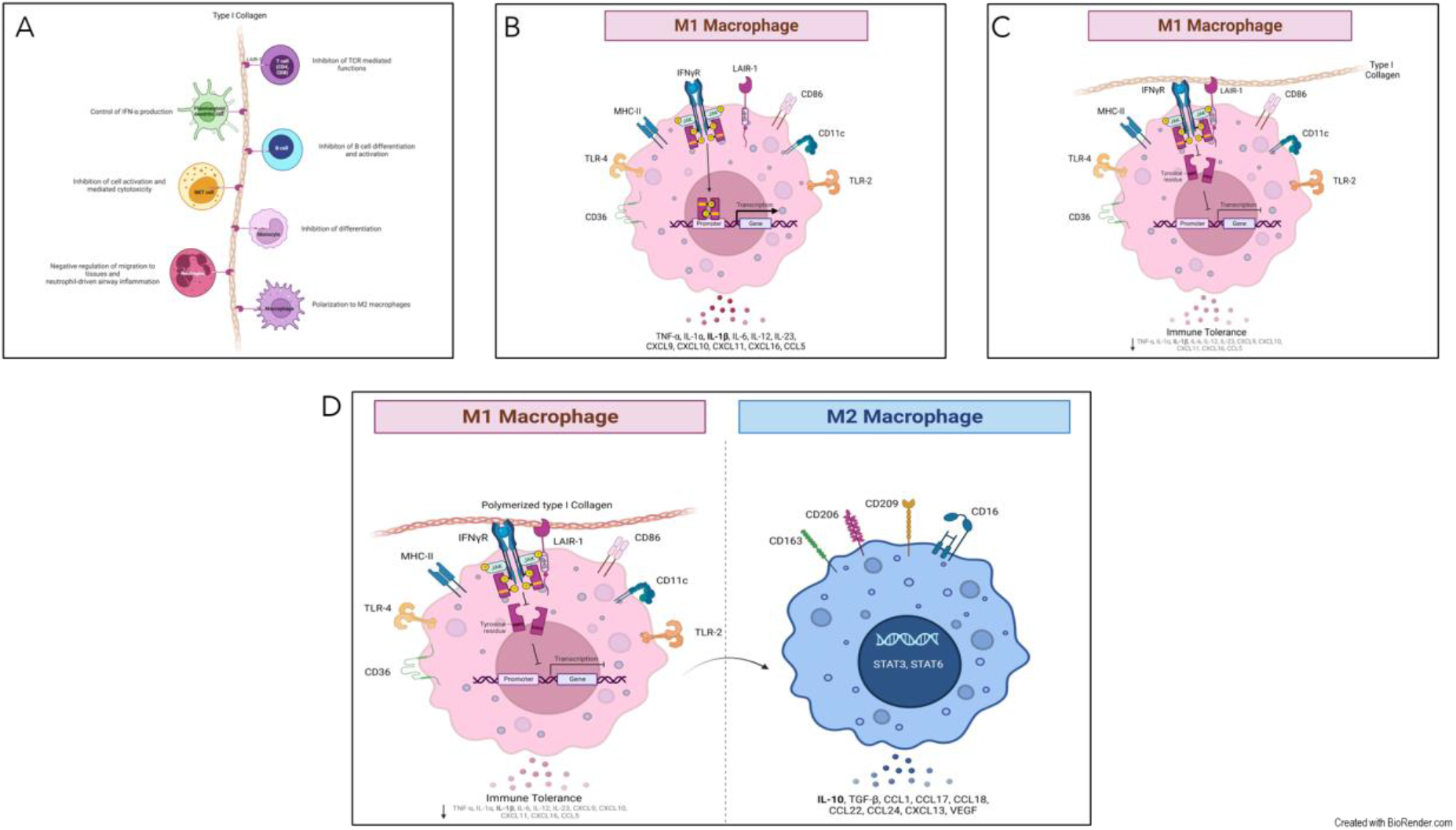
PTIC regulates STAT-1 phosphorylation through LAR-1 in M1 macrophages and favors polarization towards M2 macrophages. (A) Leukocyte-associated immunoglobulin-like receptor 1 (LAIR1, or CD305) is a type I transmembrane glycoprotein that contains one extracellular Ig-like domain and two immunoreceptor tyrosine-based inhibitory motifs (ITIMs) in its intracellular domain. LAIR1 is expressed in most hematopoietic lineages, including monocytes, macrophages, dendritic cells (DCs), natural killer (NK) cells, and many T and B cell populations. Its extracellular domain binds to glycine-proline-hydroxyproline collagen repeats, and its ITIMs recruit phosphatases SHP-1 and SHP-2. Collagens, C1q, MBL, surface protein-D (SP-D), Rifins, and Colec12, have been reported as ligands for LAIR1. It downregulates T, B, and natural killer (NK) cell functions by recruitment of SHP1 and SHP2 phosphatases. (B) The pre-polarized (M0-) macrophage subsets challenge with LPS/IFN-γ induce polarization to M1-macrophages. The human monocytic cell line THP-1 expresses high levels of LAIR1. (C) LAIR1 binding type I collagen regulates immune system balance and protects against tissue damage against a hyperactive immune response or autoimmune dysfunction through SHP-1, SHP-2, CSK, and pSTAT-1 intracellular signaling. (D) Polymerized type I collagen induces down phosphorylation of STAT-1 in M1-macrophages and promotes the polarization to M2-macrophages.

## Data Availability

All data produced in the present study are available upon reasonable request to the authors

## ACKNOWLEDGMENTS

Polymerized type I collagen and type I collagen were donated by Aspid SA de CV.

## AUTHOR CONTRIBUTIONS

Furuzawa-Carballeda and Torres-Villalobos had full access to all of the data in the study and take responsibility for the integrity of the data in the accuracy of the data analysis.

*Concept and design:* Furuzawa-Carballeda

*Acquisition, analysis, or interpretation of data:* Méndez-Flores, Priego-Ranero, Azamar-Llamas, Olvera-Prado, Rivas-Redondo, Ochoa-Hein, Urbina-Terán, Septién-Stute, Hernández-Gilsoul, Olivares-Martínez, and Hernández-Ramírez.

Drafting of the manuscript Olivares-Martínez, Hernández-Ramírez; Núñez-Álvarez, Furuzawa-Carballeda, and Torres-Villalobos.

*Critical revision of the manuscript for important intellectual content:* Chapa, Méndez-Flores, Priego-Ranero, Azamar-Llamas, Olvera-Prado, Rivas-Redonda, Ochoa-Hein, López- Mosqueda, Rojas-Castañeda, Urbina-Terán, Septién-Stute, Hernández-Gilsoul,Diana Aguilar-León, Méndez-Flores, Priego-Ranero, Azamar-Llamas, Olvera-Prado, Rivas- Redondo, Ochoa-Hein, Rojas-Castañeda, Olivares-Martínez, Hernández-Ramírez, Aguilar-León, Furuzawa-Carballeda and Torres-Villalobos.

*Statistical analysis:* Olivares-Martínez, Hernández-Ramírez and Nuñéz-Álvarez.

*Obtained funding:* Furuzawa-Carballeda

*Supervision:* Furuzawa-Carballeda, and Torres-Villalobos

## AVAILABILITY OF DATA AND MATERIALS

Data available on request from the authors

## ETHICS APPROVAL AND CONSENT TO PARTICIPATE

The study was approved by the institutional review board at Instituto Nacional de Ciencias Médicas y Nutrición Salvador Zubirán (INCMNSZ, reference no. IRE 3412-20-21-1) and was conducted in compliance with the Declaration of Helsinki (World Medical Association. World Medical Association Declaration of Helsinki. *JAMA*. 2013;310(20):2191-2194.), the Good Clinical Practice guidelines, and local regulatory requirements. All participants provided written informed consent.

## ROLE OF THE FOUNDING SOURCE

The funder of the study had no role in the study design, data collection, data analysis, data interpretation, or writing of the report. The corresponding authors had full access to all the data in the study and had final responsibility for the decision to submit for publication.

